# 5-hydroxymethylcytosine Epigenetic Markers in COVID-19-Associated Acute Coronary Syndrome: Insights into Neutrophil Activation and PDE4D Upregulation

**DOI:** 10.1101/2024.09.09.24313367

**Authors:** Zhongheng Li, Maimaitiyasen Duolikun, Hangyu Chen, Lei Zhang, Yishuo Liu, Ruining Li, Dan Li, Lijie Sun, Long Chen

**Affiliations:** Department of Cardiology, Peking University Third Hospital, NHC Key Laboratory of Cardiovascular Molecular Biology and Regulatory Peptides, Beijing 100191, China; Department of Pharmacy, Peking University Third Hospital, Beijing 100191, China; Peking University Health Science Center, Beijing 100191, China; Key Laboratory of Tropical Biological Resources of Ministry of Education, School of Pharmaceutical Sciences, Hainan University, Haikou 570100, China

**Keywords:** COVID19, Acute Coronary Syndrome, 5-Hydroxymethylcytosine, cell-free DNA, PDE4D

## Abstract

**Background:** Studies have reported that 5hmC features in cell-free DNA (cfDNA) could serve as early warning biomarkers for the occurrence and progression of COVID-19, as well as myocardial injury. However, its roles in the occurrence and progression of acute coronary syndrome (ACS) following COVID-19 infection have not been fully studied.

**Methods:** Firstly, we used the 5hmC-Seal technique to obtain genome-wide 5hmC profiles from plasma cfDNA of 24 ACS2N patients (individuals experiencing ACS onset within 2 months after COVID-19 infection), 28 ACS2W patients (individuals experiencing ACS onset beyond 2 months after COVID-19 infection), and 16 ACS patients (patients with ACS without COVID-19 infection). Secondly, we performed GO, KEGG analysis on the differentially expressed genes and identified a series of immune and inflammation related genes. Thirdly, the distribution of immune cells in different groups of patients was studied by immune infiltration analysis. Finally, we performed PPI network analysis on these genes to identify potential key target genes.

**Results:** In this study, we firstly found that there was a significant difference in 5hmC levels between ACS2N patients and ACS patients, while the difference between ACS2W and ACS was not significant. Secondly, it was found that neutrophils were abnormally activated in the ACS2N group. Finally, a target gene phosphodiesterase 4D (PDE4D) was found to be highly expressed in the ACS2N group by PPI network analysis of the differential genes and validated with external datasets.

**Conclusions:** Our study suggested that 5hmC markers extracted from plasma cfDNA could differentiate between ACS2N and ACS patients. In addition, we observed that neutrophils exhibited abnormal activation in ACS2N patients. Further analysis showed that COVID-19 infection may affect the occurrence and development of ACS by abnormally up-regulating PDE4D gene expression.

## INTRODUCTION

In the global pandemic of coronavirus disease 2019 (COVID-19), subsequent studies have found that COVID-19 not only impacts the care and outcomes of ACS patients in general, but also affects the incidence and outcomes of ACS itself. A UK study compared 156 ACS patients with COVID-19 to 6,708 ACS patients without COVID-19, revealing a three-fold increase in mortality among those with COVID-19[1]. Another retrospective study matched 62 ST segment Elevation Myocardial Infarction(STEMI)patients with COVID-19 to 310 non-COVID-19 STEMI patients, and found a higher in-hospital mortality rate (29% vs. 5.5%), a five-fold increase in in-stent thrombosis, and a two-fold increase in heart failure rates[2]. In addition to acute phase risks, Xie et al.[3] compared approximately 153,000 survivors of COVID-19, and found a Hazard Ratio (HR) of 1.72 for ACS incidence within 12 months post-infection. Given the significant health burdens of cardiovascular events, an in-depth exploration of the mechanisms underlying COVID-19-induced ACS in both the near and long term is essential to guide future treatment and prevention efforts. Therefore, there is an urgent need to reveal these complex molecular mechanisms through early screening biomarkers and to adopt effective interventions. This is not only key to reducing the incidence of ACS, but also an important strategy to reduce the risk of complications and improve the short- and long-term prognosis of COVID-19 patients.

Mechanisms contributing to the poor prognosis of patients with COVID-19-positive ACS may include severe endothelial dysfunction, which may be caused by direct viral invasion of endothelial cells, activation of the immune response, cytokine storms, hypoxemia, severe infections, changes in shear stress, or activation of the renin-angiotensin-aldosterone system (RAAS)[4]. Furthermore, inflammatory mechanisms induced by COVID-19 may trigger plaque rupture and create a pro-thrombotic environment, potentially increasing the risk of thrombosis and impairing reperfusion, thereby increasing the risk of in-stent thrombosis[5]. Studies have shown that even after the acute phase, the risk of new-onset cardiovascular disease increases regardless of prior cardiovascular risk factors[3]. Although we have a preliminary understanding of the molecular mechanisms of COVID-19-induced cardiovascular complications[6], much still remains unknown. Such as changes in gene level, the much earlier stage of the disease, etc.

Epigenetics, being an area of research that explores gene-environment interactions, has gradually gained importance in the study of atherosclerosis and other cardiovascular, diseases in recent years[7, 8]. DNA hydroxymethylation is a type of epigenetic modification, a relatively stable derivative produced by the demethylation of 5-methylcytosine (5mC), mediated by the TET (ten-eleven translocation) family of proteins[9]. As circulating cell-free DNA (cfDNA) has the potential to provide liquid biopsy-based noninvasive assays and methods for human disease assessment, technological advances have generated great interest in identifying 5-hydroxymethylcytosine (5hmC) modifications in cfDNA[10]. In patients with ST segment Elevation Myocardial Infarction (STEMI), researchers identify strong cardiac cfDNA signaling that correlates with the elevation-attenuation dynamics of troponin and creatine phosphokinase after coronary angioplasty[11]. In addition, decreases in the expression of TET2 and 5hmC were positively associated with vascular injury and severity of atherosclerotic disease[12]. However, the modification profile of 5hmC in peripheral blood cfDNA of different patients who developed ACS after contracting COVID-19 is still unknown, and whether 5hmC modification is involved in the progression of ACS triggered by neocoronary and its potential as a potential biomarker for noninvasive monitoring of ACS needs to be further investigated and widely explored. Therefore, 5hmC in cfDNA has the potential to be developed as an earlier biomarker for ACS patients following COVID-19 infection.

In this study, based on the varying clinical presentations of patients, we hypothesize that there are differences in cfDNA methylation among ACS patients who have had COVID-19 infection within the past two months. Our aim is to identify different mechanisms of COVID-19-induced ACS through differential gene methylation expression profiling, thus providing insights for intervention and prognosis of these ACS patients.

## Materials and methods

### Participants

The study enrolled patients aged 18 years and older with complete medical history, clinical, and biochemical indicator information, who were hospitalized in the Coronary Care Unit of Peking University Third Hospital for treatment of ACS from January 2023 to March 2023. Specifically, we aimed to compare patients who experienced ACS for the first time after COVID-19 infection with those who experienced ACS for the first time during the same period but had not got COVID-19 infection. Inclusion criteria included Patients diagnosed with acute coronary syndrome upon hospital admission between 18 and 80 years old; and willing to sign informed consent and cooperate with follow-up as well. Exclusion criteria include pregnancy, chronic kidney disease (estimated glomerular filtration rate < 60 ml/min), acute or chronic liver disease, malignancy, chronic obstructive pulmonary disease or pulmonary heart disease, severe systemic diseases (such as septic shock, amyloidosis, systemic lupus erythematosus, vasculitis, thrombocytopenic purpura, and etc.), specific infectious diseases (such as viral hemorrhagic fever, hepatitis C, and etc.), self-discharge from hospital.

According to the temporal relationship between ACS onset and COVID-19 infection, patients were categorized into three groups: ACS within 2 months after COVID-19 infection group (ACS2N), ACS more than 2 months after COVID-19 infection group (ACS2W), and ACS group without COVID-19 infection (ACS), including 26, 29, and 16 patients respectively. All patients provided informed consent. The diagnosis of ACS and COVID-19 in all cases was made using standardized diagnostic criteria. The study was conducted in compliance with the Declaration of Helsinki.

### Collection and Analysis of Clinical Samples, along with Isolation of cfDNA from peripheral blood

Blood samples (8-10 mL) were obtained from all patients via routine intravenous blood sampling and collected into a Cell-Free DNA collection tube (Roche). The metadata included demographic characteristics (age, gender, body mass index (BMI), smoking, diabetes mellitus, hyperlipemia, hypertension and so on.); laboratory characteristics(hemoglobin, HbA1c, hypersensitive c-reactive protein (hsCRP), urea, creatinine (Cr), aspartate aminotransferase (AST), alanine aminotransferase (ALT), cholesterol (Chol), triglyceride (TG), high-density lipoprotein (HDL),low-density lipoprotein (LDL), peak value of 24h creatine kinase-MB (CK-MB), peak value of troponin T (TnT) and so on), echocardiography (myocardial segment, left ventricular ejection fractions (LVEF) and so on), classification of acute coronary syndromes, follow-up results (length of hospital stay, major adverse cardiovascular events (MACE) occurred within 12 months).

MACE is typically defined as including at least one of the following events: cardiovascular death: death due to cardiovascular causes such as myocardial infarction, arrhythmias, etc.; non-fatal myocardial infarction: occurrence of myocardial infarction without resulting in death; unplanned coronary revascularization: non-scheduled coronary artery intervention or stenting typically due to acute coronary syndrome (e.g., angina, myocardial infarction); non-fatal stroke: Ischemic stroke or transient ischemic attack.

Plasma separation was performed within 24 hours, with whole blood samples being centrifuged twice at 4°C, first at 1350×g for 12 minutes and then at 13,500×g for another 12 minutes. The plasma layers were subsequently transferred to a new tube and immediately stored at −80℃ for future use. The plasma cfDNA was extracted from 2-4 mL of plasma using the Quick-cfDNA Serum & Plasma Kit (ZYMO) and then stored at −80℃. Prior to library preparation, the concentration and quality of cfDNA were quantified using a Qubit fluorometer and nucleic acid electrophoresis. Follow-up methods primarily include telephone follow-ups and electronic health record monitoring.

### 5hmC library construction and high-throughput sequencing

5hmC libraries for all cfDNA samples were constructed using the high-efficiency hmC-Seal technology described previously[13]. Briefly, 1-10ng of cfDNA were end repaired, followed by 3’-adenylation using a Hyper Prep kit (KAPA Biosystems) and ligated with Illumina-compatible adapters on both ends. Then, the ligated DNA was incubated in a 25 μL solution containing 25 mM MgCl2, 50 mM HEPES buffer (pH=8.0), 100 µM UDP-6-N3-Glc and 1 µM β -glucosyltransferase for 2 h at 37°C. After DNA purified by the DNA Clean & Concentrator Kit (ZYMO), DBCO-PEG4-Biotin (1 µL; Click Chemistry Tools; 4.5 mM stock in dimethyl sulfoxide) was added and incubated at 37°C for 2 h. After 2h, DNA purification was performed again with the ZYMO kit described above. Subsequently, the biotin-labeled DNA was pulled down by Streptavidin beads (2.5 µL; Life Technologies) for 30 min at room temperature. The beads were transferred to the amplification reaction after being washed. Afterwards, DNA fragments containing the 5hmC features were amplified by PCR using the Nextera DNA Sample Preparation kit. The amplified products were purified using AMPure XP beads (Beckman) according to the manufacturer’s instructions. The libraries were quantified by a Qubit fluorometer (Life Technologies). Finally, we used the Agilent 2100 bioanalyzer for quality control of the 5hmC library. Libraries that meet the quantification standards are then subjected to paired-end 150 bp high-throughput sequencing on the NovaSeq 6000 platform.

### Read mapping and 5hmC peak identification

FastQC (version 0.11.5) was used to check the sequence quality. Paired-end reads were initially processed using Trim_Galore (https://github.com/FelixKrueger/TrimGalore) [14] to eliminate adapter sequences and low-quality nucleotides. Acquired high-quality reads data were compared with the human reference genome (hg19/GRCh37) using Bowtie2 v2.2.9 [15] and the PCR duplicates were depleted by Samtools software [16]. After the extension process, we transformed the paired-end reads into BedGraph format. Subsequently, we normalized these reads to the total count of aligned reads, utilizing the bedtools software package, version 2.19.1 [17]. Meanwhile, leveraging the UCSC Genome Browser’s BedGraphToBigWig utility, we transformed the paired-end reads into BigWig format. This conversion facilitated the visualization process, enabling the use of the Integrated Genomics Viewer for a comprehensive analysis.

### Identification of the DEGs

Here, we performed differential expression analysis using the R Bioconductor software package limma (V3.46.0) to identify differentially expressed genes [18]. Following grouping of the samples, identification of differentially expressed genes between the ACS2N group and the ACS2W group and between the ACS2N group and the ACS group was performed using DESeq2 and deeptools (|log2FC| > 0.4 and p-value<0.05). Volcano plots were made by the R software ggplot2 V3.3.5 package, and VennDiagram was drawn with the R software VennDiagram V1.6.20 package [19]. Heat maps of the 399 DEGs obtained by taking the intersection of the two groups were drawn by the R software Pheatmap V1.0.12 package.

### Functional enrichment analysis of DEGs

To explore the function and pathway of the overlapping DEGs, the functional enrichment analysis was performed in metascape database(Metascape)((significant as a P<0.05 and a q-value < 0.05). For all overlapping DEGs, gene ontology (GO) terms (BP, biological process) as well as Kyoto encyclopedia of genes and genomes (KEGG) pathways enrichment analysis were conducted and visualized.

### Analysis of immune cell infiltration of DEGs

CIBERSORT was then performed to analyze the infiltration of 22 kinds of the immune cells. We obtained the relative abundance of infiltrated immune cell according to P<0.05, and then drew the correlation heatmap for visualizing the correlation of infiltrated immune cells through R software corrplot V0.92 package, and next explored the differential infiltration in immune cells between atherosclerosis and control groups using Wilcoxon rank sum test, and subsequently analyzed the Spearman relationship between biomarkers and infiltrating immune cells. The results was visualized via R software ggstatsplot V0.9.1 package.

### Single-cell RNA sequencing data analysis

The barcodes data, gene features data, and gene count matrix data of GSE235436 preprocessed by Cellranger (10X Genomics) were downloaded from GEO database. These data were imported in R, and analyzed using Seurat V4.1.0 package [20]. Firstly, quality control was conducted through filtering out cells satisfying the following criteria: a gene count per cell > 200 and < 2500, and a percentage of mitochondrial genes < 5%. Next, the data were normalized by NormalizeData function. For the downstream analysis, top-ranked 2000 variably expressed genes were selected using “vst” method in FindVariableFeatures function. Before the PCA, the data were scaled using ScaleData function. The data were then subjected to PCA, cluster analysis, and Uniform Manifold Approximation and Projection (UMAP) dimensional reduction with RunPCA, FindClusters, and Run UMAP functions. Subsequently, the cell clusters were visualized using the UMAP plots displayed by the DimPlot function. Different expressions of signatures were determined with the FindAllMarkers function. Violin plots were drawn using the VlnPlot function. Furthermore, we applied R software SingleR V1.4.1 package[21] to annotate cell types, and we employed the celldex V1.0.0 package to download the HumanPrimaryCellAtlasData reference.

### Construction of PPI networks and prediction of key genes

In this study, we utilized the STRING online database to construct a protein-protein interaction (PPI) network for differentially expressed genes, setting the interaction score threshold to >0.4. Subsequently, we employed the CytoHubba plugin to identify significant genes within the PPI network, designating them as key hub genes [22].

### Statistical analysis

Statistical analysis was performed using GraphPad Prism 8.0.2. Data between groups were compared using One-way ANOVA and the two groups of data were compared by t test. Measurement data obtained from the experiment were expressed as means ± standard and P <0.05 was considered to be statistically significant.

## RESULTS

### Clinical characteristics of ACS2N, ACS2W and ACS patients

Plasma samples from 26 ACS2N patients, 29 ACS2W patients, and 16 ACS patients were collected. Clinical data were collected from all samples, and detailed information is listed in **Table1**. Among the three groups, significant differences were detected in Hemoglobin (P=0.027), HbA1c (P=0.025). No significant differences were found in demographic characteristics (Age, Gender, BMI, Smoking, Diabetes mellitus, Hyperlipemia, Hypertension.), Laboratory characteristics (hsCRP, Urea, Cr, AST, ALT, Chol, TG, HDL,LDL, Peak value of 24h CK-MB, Peak value of TnT), Echocardiography (Myocardial segment, LVEF), Classification of acute coronary syndromes, Follow-up results (Length of hospital stay, MACE occurred within 12 months). Although the data show that among the three groups, the ACS2N group had a higher incidence of MACE events and hospitalization days within 12 months, these differences were not statistically significant.

**TABLE1.**
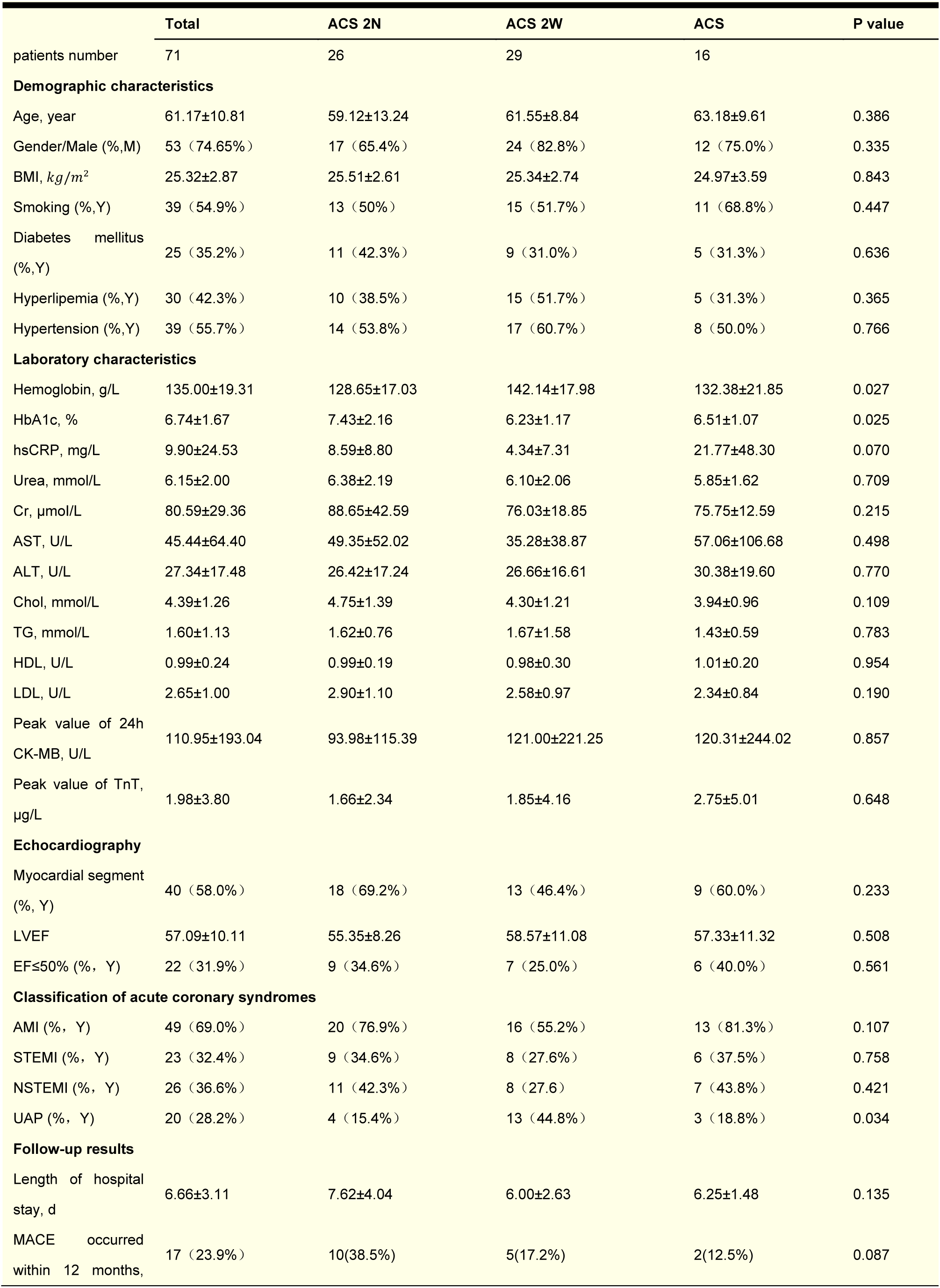
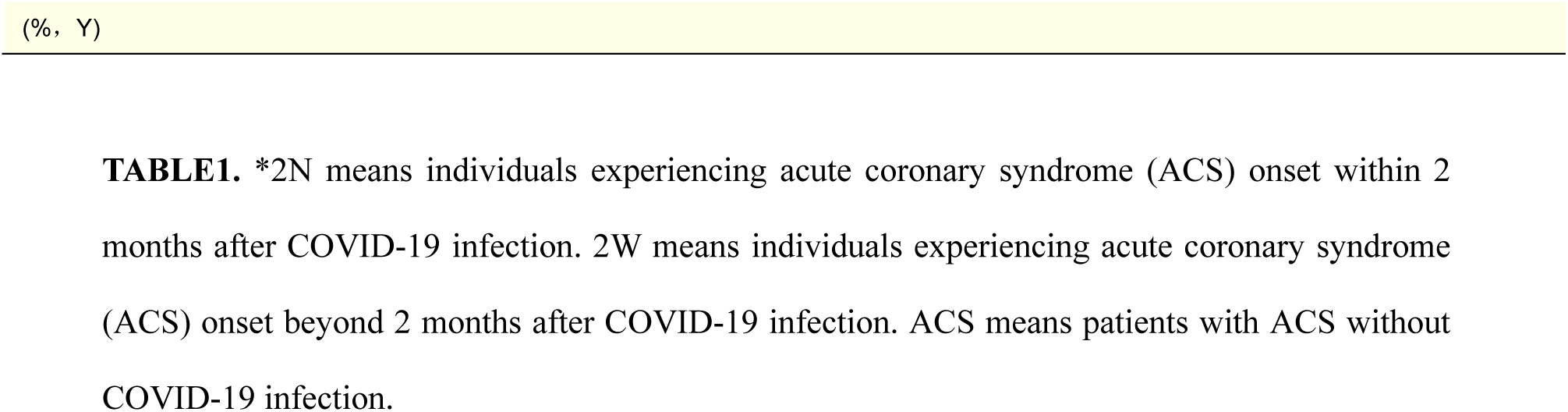
The summary of basic characteristics and clinically relevant indices for all participating patients in this study.

\*2N means individuals experiencing acute coronary syndrome (ACS) onset within 2 months after COVID-19 infection. 2W means individuals experiencing acute coronary syndrome (ACS) onset beyond 2 months after COVID-19 infection. ACS means patients with ACS without COVID-19 infection.
|  | Total | ACS 2N | ACS 2W | ACS | P value |
| --- | --- | --- | --- | --- | --- |
| patients number | 71 | 26 | 29 | 16 |  |
| **Demographic characteristics** |  |  |  |  |  |
| Age, year | 61.17±10.81 | 59.12±13.24 | 61.55±8.84 | 63.18±9.61 | 0.386 |
| Gender/Male (% ,M) | 53 (74.65%) | 17 (65.4%) | 24 (82.8%) | 12 (75.0%) | 0.335 |
| BMI, $kg/m^2$ | 25.32±2.87 | 25.51±2.61 | 25.34±2.74 | 24.97±3.59 | 0.843 |
| Smoking (% ,Y) | 39 (54.9%) | 13 (50%) | 15 (51.7%) | 11 (68.8%) | 0.447 |
| Diabetes mellitus (% ,Y) | 25 (35.2%) | 11 (42.3%) | 9 (31.0%) | 5 (31.3%) | 0.636 |
| Hyperlipemia (% ,Y) | 30 (42.3%) | 10 (38.5%) | 15 (51.7%) | 5 (31.3%) | 0.365 |
| Hypertension (% ,Y) | 39 (55.7%) | 14 (53.8%) | 17 (60.7%) | 8 (50.0%) | 0.766 |
| **Laboratory characteristics** |  |  |  |  |  |
| Hemoglobin, g/L | 135.00±19.31 | 128.65±17.03 | 142.14±17.98 | 132.38±21.85 | 0.027 |
| HbA1c, % | 6.74±1.67 | 7.43±2.16 | 6.23±1.17 | 6.51±1.07 | 0.025 |
| hsCRP, mg/L | 9.90±24.53 | 8.59±8.80 | 4.34±7.31 | 21.77±48.30 | 0.070 |
| Urea, mmol/L | 6.15±2.00 | 6.38±2.19 | 6.10±2.06 | 5.85±1.62 | 0.709 |
| Cr, μmol/L | 80.59±29.36 | 88.65±42.59 | 76.03±18.85 | 75.75±12.59 | 0.215 |
| AST, U/L | 45.44±64.40 | 49.35±52.02 | 35.28±38.87 | 57.06±106.68 | 0.498 |
| ALT, U/L | 27.34±17.48 | 26.42±17.24 | 26.66±16.61 | 30.38±19.60 | 0.770 |
| Chol, mmol/L | 4.39±1.26 | 4.75±1.39 | 4.30±1.21 | 3.94±0.96 | 0.109 |
| TG, mmol/L | 1.60±1.13 | 1.62±0.76 | 1.67±1.58 | 1.43±0.59 | 0.783 |
| HDL, U/L | 0.99±0.24 | 0.99±0.19 | 0.98±0.30 | 1.01±0.20 | 0.954 |
| LDL, U/L | 2.65±1.00 | 2.90±1.10 | 2.58±0.97 | 2.34±0.84 | 0.190 |
| Peak value of 24h CK-MB, U/L | 110.95±193.04 | 93.98±115.39 | 121.00±221.25 | 120.31±244.02 | 0.857 |
| Peak value of TnT, μg/L | 1.98±3.80 | 1.66±2.34 | 1.85±4.16 | 2.75±5.01 | 0.648 |
| **Echocardiography** |  |  |  |  |  |
| Myocardial segment (% , Y) | 40 (58.0%) | 18 (69.2%) | 13 (46.4%) | 9 (60.0%) | 0.233 |
| LVEF | 57.09±10.11 | 55.35±8.26 | 58.57±11.08 | 57.33±11.32 | 0.508 |
| EF≤50% (% , Y) | 22 (31.9%) | 9 (34.6%) | 7 (25.0%) | 6 (40.0%) | 0.561 |
| **Classification of acute coronary syndromes** |  |  |  |  |  |
| AMI (% , Y) | 49 (69.0%) | 20 (76.9%) | 16 (55.2%) | 13 (81.3%) | 0.107 |
| STEMI (% , Y) | 23 (32.4%) | 9 (34.6%) | 8 (27.6%) | 6 (37.5%) | 0.758 |
| NSTEMI (% , Y) | 26 (36.6%) | 11 (42.3%) | 8 (27.6%) | 7 (43.8%) | 0.421 |
| UAP (% , Y) | 20 (28.2%) | 4 (15.4%) | 13 (44.8%) | 3 (18.8%) | 0.034 |
| **Follow-up results** |  |  |  |  |  |
| Length of hospital stay, d | 6.66±3.11 | 7.62±4.04 | 6.00±2.63 | 6.25±1.48 | 0.135 |
| MACE occurred within 12 months, | 17 (23.9%) | 10(38.5%) | 5(17.2%) | 2(12.5%) | 0.087 |

|  | Total | ACS 2N | ACS 2W | ACS | P value |
| --- | --- | --- | --- | --- | --- |
| patients number | 71 | 26 | 29 | 16 |  |
| **Demographic characteristics** |  |  |  |  |  |
| Age, year | 61.17±10.81 | 59.12±13.24 | 61.55±8.84 | 63.18±9.61 | 0.386 |
| Gender/Male (% ,M) | 53 (74.65%) | 17 (65.4%) | 24 (82.8%) | 12 (75.0%) | 0.335 |
| BMI, kg/m<sup>2</sup> | 25.32±2.87 | 25.51±2.61 | 25.34±2.74 | 24.97±3.59 | 0.843 |
| Smoking (% ,Y) | 39 (54.9%) | 13 (50%) | 15 (51.7%) | 11 (68.8%) | 0.447 |
| Diabetes mellitus (% ,Y) | 25 (35.2%) | 11 (42.3%) | 9 (31.0%) | 5 (31.3%) | 0.636 |
| Hyperlipemia (% ,Y) | 30 (42.3%) | 10 (38.5%) | 15 (51.7%) | 5 (31.3%) | 0.365 |
| Hypertension (% ,Y) | 39 (55.7%) | 14 (53.8%) | 17 (60.7%) | 8 (50.0%) | 0.766 |
| **Laboratory characteristics** |  |  |  |  |  |
| Hemoglobin, g/L | 135.00±19.31 | 128.65±17.03 | 142.14±17.98 | 132.38±21.85 | 0.027 |
| HbA1c, % | 6.74±1.67 | 7.43±2.16 | 6.23±1.17 | 6.51±1.07 | 0.025 |
| hsCRP, mg/L | 9.90±24.53 | 8.59±8.80 | 4.34±7.31 | 21.77±48.30 | 0.070 |
| Urea, mmol/L | 6.15±2.00 | 6.38±2.19 | 6.10±2.06 | 5.85±1.62 | 0.709 |
| Cr, µmol/L | 80.59±29.36 | 88.65±42.59 | 76.03±18.85 | 75.75±12.59 | 0.215 |
| AST, U/L | 45.44±64.40 | 49.35±52.02 | 35.28±38.87 | 57.06±106.68 | 0.498 |
| ALT, U/L | 27.34±17.48 | 26.42±17.24 | 26.66±16.61 | 30.38±19.60 | 0.770 |
| Chol, mmol/L | 4.39±1.26 | 4.75±1.39 | 4.30±1.21 | 3.94±0.96 | 0.109 |
| TG, mmol/L | 1.60±1.13 | 1.62±0.76 | 1.67±1.58 | 1.43±0.59 | 0.783 |
| HDL, U/L | 0.99±0.24 | 0.99±0.19 | 0.98±0.30 | 1.01±0.20 | 0.954 |
| LDL, U/L | 2.65±1.00 | 2.90±1.10 | 2.58±0.97 | 2.34±0.84 | 0.190 |
| Peak value of 24h CK-MB, U/L | 110.95±193.04 | 93.98±115.39 | 121.00±221.25 | 120.31±244.02 | 0.857 |
| Peak value of TnT, µg/L | 1.98±3.80 | 1.66±2.34 | 1.85±4.16 | 2.75±5.01 | 0.648 |
| **Echocardiography** |  |  |  |  |  |
| Myocardial segment (% , Y) | 40 (58.0%) | 18 (69.2%) | 13 (46.4%) | 9 (60.0%) | 0.233 |
| LVEF | 57.09±10.11 | 55.35±8.26 | 58.57±11.08 | 57.33±11.32 | 0.508 |
| EF≤50% (% , Y) | 22 (31.9%) | 9 (34.6%) | 7 (25.0%) | 6 (40.0%) | 0.561 |
| **Classification of acute coronary syndromes** |  |  |  |  |  |
| AMI (% , Y) | 49 (69.0%) | 20 (76.9%) | 16 (55.2%) | 13 (81.3%) | 0.107 |
| STEMI (% , Y) | 23 (32.4%) | 9 (34.6%) | 8 (27.6%) | 6 (37.5%) | 0.758 |
| NSTEMI (% , Y) | 26 (36.6%) | 11 (42.3%) | 8 (27.6%) | 7 (43.8%) | 0.421 |
| UAP (% , Y) | 20 (28.2%) | 4 (15.4%) | 13 (44.8%) | 3 (18.8%) | 0.034 |
| **Follow-up results** |  |  |  |  |  |
| Length of hospital stay, d | 6.66±3.11 | 7.62±4.04 | 6.00±2.63 | 6.25±1.48 | 0.135 |
| MACE occurred within 12 months, | 17 (23.9%) | 10(38.5%) | 5(17.2%) | 2(12.5%) | 0.087 |

### 5hmC distribution in plasma cfDNA from 3 groups of ACS patients

We used the 5hmC-Seal technique for cfDNA samples in this study, which aimed to mapping the genome-wide 5hmC distribution. Firstly, we performed a rigorous quality control (QC) analysis on samples from the ACS, ACS2N and ACS2W groups (Supplementary Figures S1A-D), and there were 2 blood samples in ACS2N group and 1 blood sample in ACS2W group that did not meet the quality inspection standards. Subsequently, sequencing data showed that the 5hmC distribution level was highest in ACS2W patients and lowest in ACS2N patients (Fig. 1A). 5hmC peaks are mainly enriched in key regulatory regions of the genome, including promoters, introns and distal regulatory regions, which is in agreement with a previous report[23], suggesting that the accumulation of 5hmC is associated with transcriptional activity (Fig. 1B). However, since no significant difference was found in the comparison between the ACS2W group and the ACS group, we proceeded to screen and analyze the differentially expressed genes in the ACS2N group compared with the above two groups separately. Visual analysis of the volcano plot revealed a total of 1,355 differentially expressed genes (DEGs) screened compared to the ACS2W group, while 2,124 DEGs were identified compared to the ACS group (Fig. 1C-D). Next, these two groups of up- and down-regulated DEGs were screened separately, and a total of 399 DEGs were screened (Fig. 1E-F, Supplementary Table1). Using the above 399 genes for clustering analysis, it can be seen that genes with similar expression within the group clustered together and appeared in the same group (Fig. 1G). Our results show that the characteristics of hydroxymethylation modification are different in ACS2N, ACS2W, and ACS patients.

**Figure 1.**
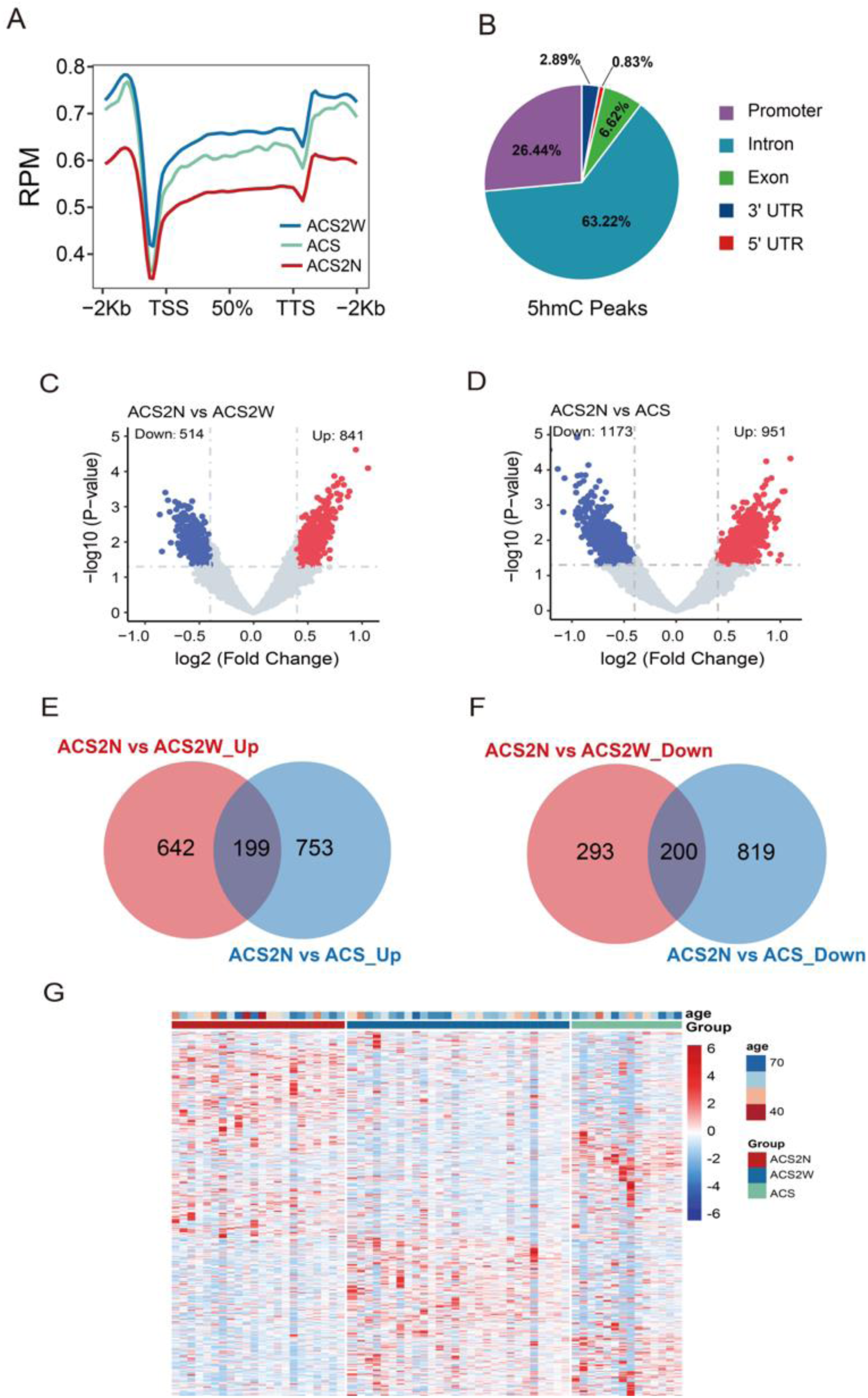
Characteristics of 5hmC distribution in plasma cfDNA of patients with ACS and patients with COVID-19 combined ACS, and screening of differentially expressed genes. A, Genome-wide distribution of 5hmC marks in plasma cfDNA. 5hmC marks are enriched in gene bodies and depleted in the flanking regions for all three groups. B, Pie chart showing the percentage of 5hmC peaks in each class of genomic regions. C-D, Volcano plot showing the DhMGs between ACS2N and ACS2W (C) or ACS (D) groups. Significantly altered DhMGs (|log2FC| > 0.4, P-value <0.05) are highlighted in red (up) or green (down), respectively. E-F, Veen diagrams of up-regulated (E) and down-regulated (F) DhMGs in ACS2N patients vs ACS2W patients versus ACS2N patients vs ACS patients. G, Heatmap showing the 399 DhMGs with sample type, age, and sex information labeled. Unsupervised hierarchical clustering was performed across genes and samples.

### GO and KEGG pathway analysis of DEGs

Next, we performed GO and KEGG pathway functional enrichment analysis on 399 DEGs. The results showed that the significantly enriched Biological Process (BP) included cyclic adenosine monophosphate (cAMP) mediated signaling, B cell differentiation, regulation of IL-2 production, cellular response to cAMP, and regulation of MAPK cascade (Fig. 2A). In the Cellular Component (CC) centrosome, actin cytoskeleton, serine/threonine protein kinase complex, protein kinase complex, and cation channel complex were the top 5 enriched items (Fig. 2A). As for Molecular Function (MF), the most enriched terms cAMP binding, GTPase regulator activity, phosphoric ester hydrolase activity, transcription factor binding, and virus receptor activity (Fig. 2A). Subsequently, the DEGs were subjected to KEGG pathway functional enrichment analysis. cAMP signaling pathway, Neurotrophin signaling pathway, Chemokine signaling pathway, PI3K-Akt signaling pathway, and cGMP-PKG signaling pathway were considered to be the most highly enriched pathways (Fig. 2B). In addition, we also noticed that PDE4D, PDE4B and other genes were mainly enriched in the cAMP signaling pathway.

**Figure 2.**
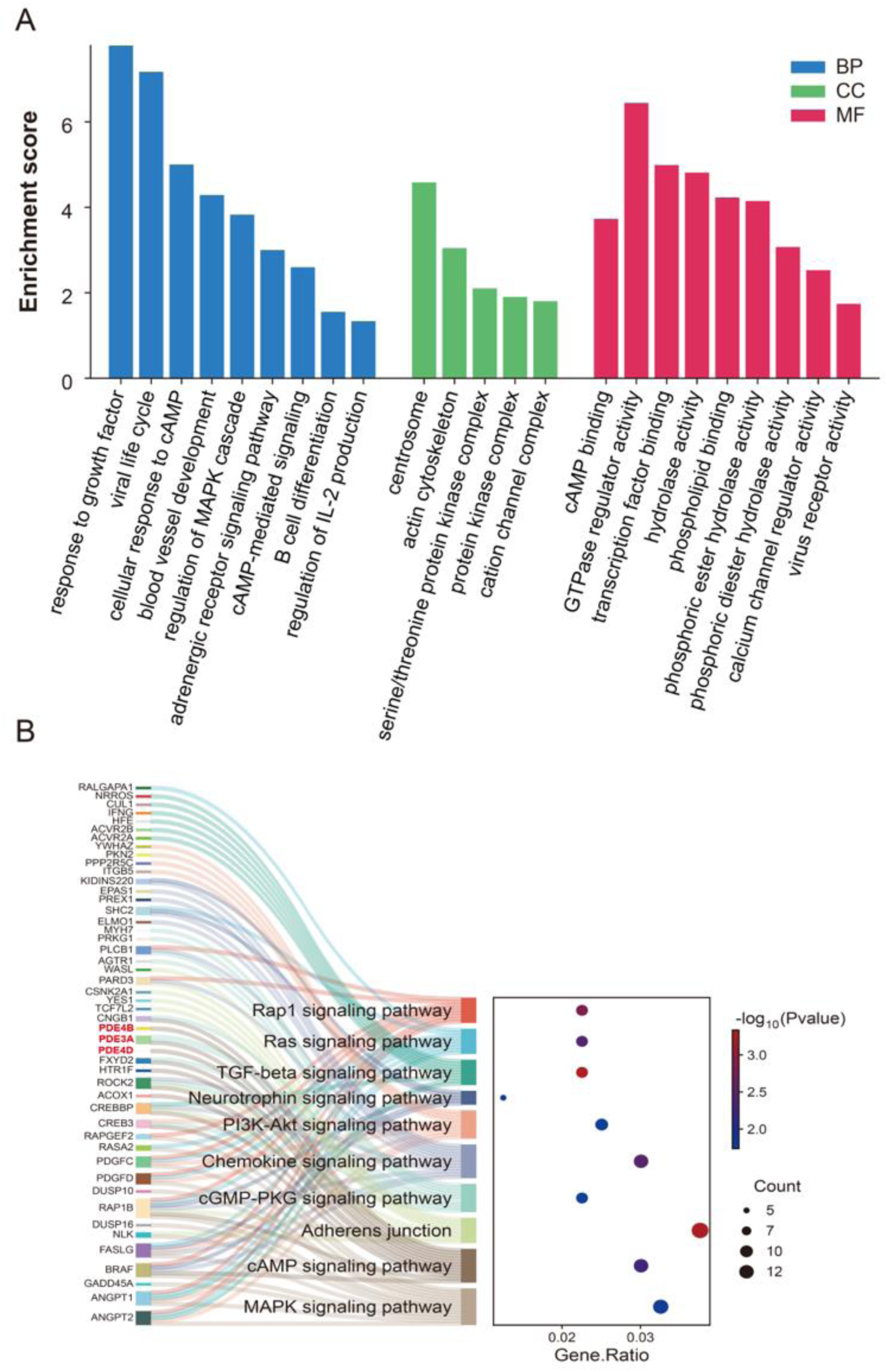
GO and KEGG pathway enrichment analysis of the 399 identified 5hmC markers by Cytoscape software. A, GO enrichment bar plot (BP: Biological Process; CC: Cellular Component; MF: Molecular Function). **B,** Plot of sankey dot KEGG pathway enrichment.

### Analysis of immune cell infiltration

We also used the CIBERSORT algorithm to analyze 22 immune cell phenotypes of 399 DEGs. As can be seen by the stacked bar graph, the percentage of neutrophils was higher in the ACS2N group (Fig. 3A). In comparison with ACS groups, ACS2N groups had a higher proportion of neutrophils, mast cells resting, T cells regulatory, and B cells naive (P<0.05). However, the T cells CD4 memory resting, NK cells resting and NK cells activates in ACS2N were relatively lower than that in ACS (all P<0.05) (Fig. 3B).

**Figure 3.**
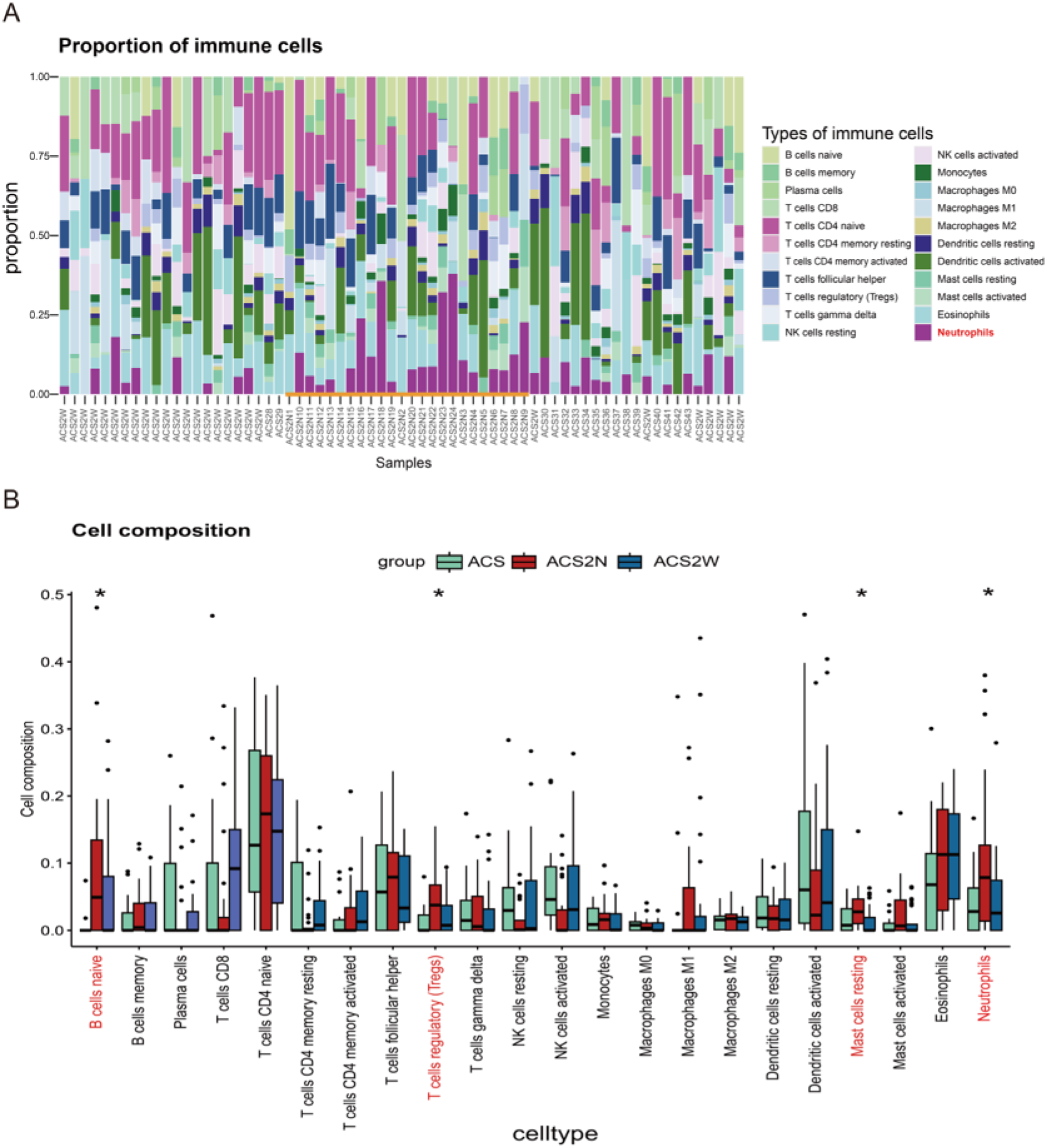
Immunoinfiltration analysis of differentially expressed genes. A, Stacked bar chart of immune cell distribution in each sample. **B,** Box-plot of the proportion of 22 types of immune cells. * P<0.05.

### Screening of target genes

Next, we performed a PPI network analysis of genes in immune and inflammation-related pathways and identified two key genes, PDE4D and PDE4B (Fig. 4A). They are highly expressed and significantly different in the ACS2N group (Fig. 4B). Subsequently, we performed a correlation type analysis of the top 5 genes with higher scores with 22 immune cells and found that PDE4D had the highest correlation with neutrophils among these genes (Fig. 4C). Finally, we targeted inflammation to PDE4D by single-cell RNA sequencing data and found that PDE4D was highly expressed in neutrophils in the disease group, which was consistent with the previous results (Fig. 4C).

**Figure 4.**
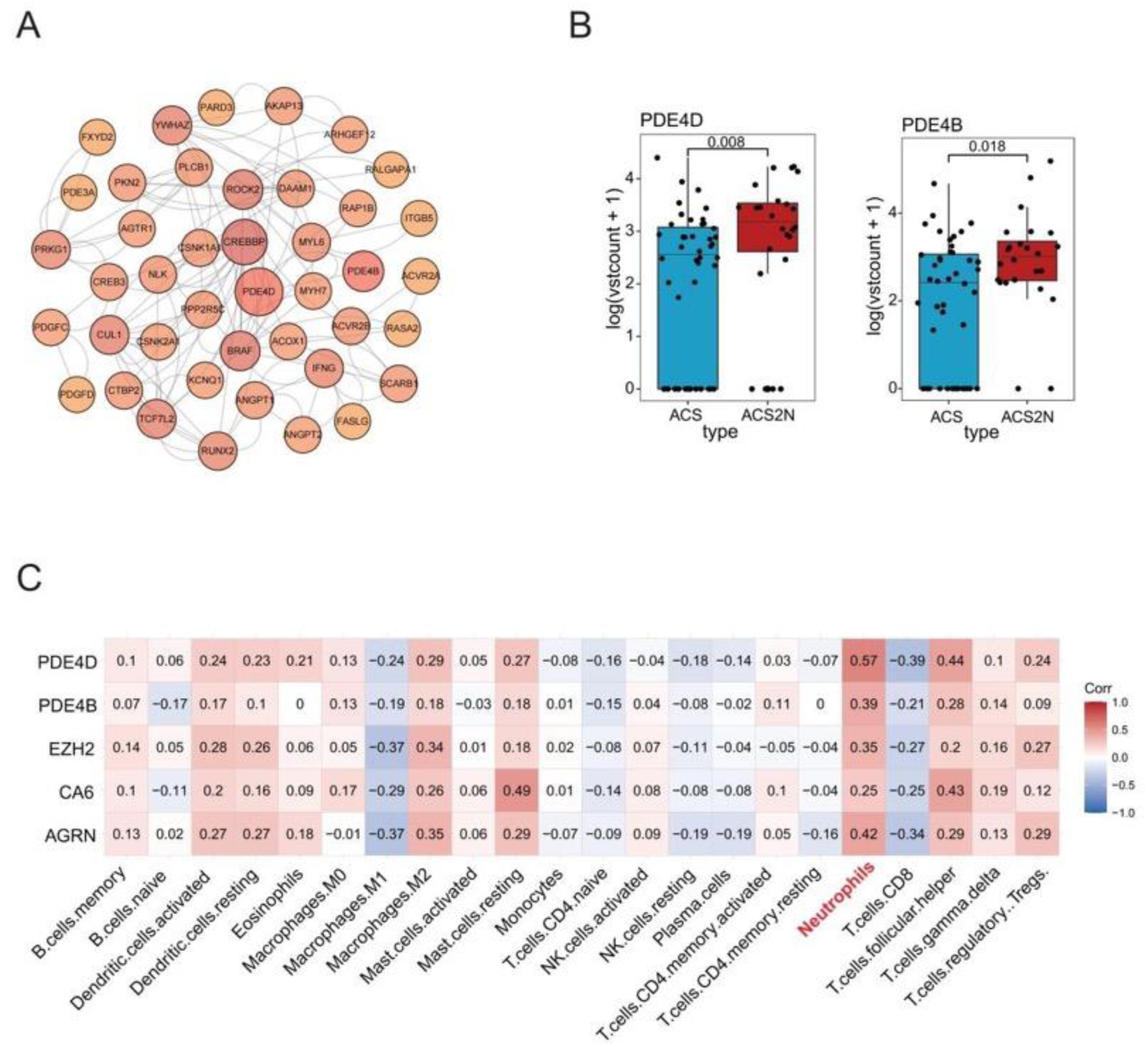
Analysis of differentially expressed genes in neutrophil-related pathways. **A,** PPI network plot of 42 genes. **B,** Boxplots of PDE4D and PDE4B. **C,** Correlations between PDE4D, PDE4B, EZH2, CA6, AGRN, and infiltrating immune cells.

## Discussion

Numerous studies consistently indicate that COVID-19 infection may significantly increase the risk of ACS and worsen prognosis[24]. In our clinical data, we particularly observed an increased incidence of segmental wall abnormalities in ACS patients within 2 months after COVID-19 infection, along with prolonged average hospital stays. Moreover, during a 12-month follow-up, there was a rise in the frequency of MACE (ACS2N: ACS2W: ACS = 38.5%:17.2%:12.5%). Due to sample size limitation, these observations have not yet shown significant statistical differences; nonetheless, they provide valuable initial insights.

5-cytosine (5mC) methylation is a fundamental epigenetic DNA modification associated with ACS [25]. In contrast, the role of 5-hydroxymethylcytosine (5hmC) modification in ACS biology and pathophysiology remains partially understood. 5hmC markers extracted from cfDNA serve as effective epigenetic biomarkers for minimally invasive or non-invasive diagnosis of coronary artery disease (CAD) [26]. However, the development of CAD is closely linked to the progression of atherosclerotic plaques, further associating it with the occurrence and development of ACS. Therefore, we investigated whether 5hmC contributes to the onset and progression of ACS following COVID-19 infection. Indeed, the abundant distribution of 5hmC in the central nervous system[27, 28] and tumors[29, 30] reveals its potential roles in neurodevelopment and tumorigenesis. As a stable yet dynamically changing epigenetic marker, 5hmC plays crucial roles in these processes. Additionally, moderate expression levels of 5hmC in the heart and skeletal muscle further underscore its importance in tissue-specific gene expression[31, 32]. Moreover, inflammation-related 5hmC markers are closely associated with the progression of atherosclerotic plaques, indicating their dynamic regulatory role in disease development. Furthermore, preliminary studies by our group reported that 5hmC features in cfDNA could serve as early warning biomarkers for the occurrence and progression of COVID-19, as well as myocardial injury[33].

Our study found significant differences in 5hmC modification levels in plasma cfDNA between ACS2N patients and ACS patients, while differences between ACS2W patients and ACS patients were minimal, consistent with our clinical observations—that is, ACS2W patients and ACS patients showed similar clinical presentations and prognoses. This difference may be attributed to the early stage of robust viral replication and inflammatory effects observed post COVID-19 infection, which may lead to neutrophil aggregation and inflammatory responses associated with severe cases. However, as the infection progresses and immune system responses evolve, some studies suggest that the virus may attenuate in later stages, indicating reduced viral replication capability and diminished inflammatory impact of the virus. This situation may lower the subsequent risk of developing other diseases, particularly after the direct inflammatory response related to the virus weakens, providing more opportunities for gradual immune recovery and regulation[34–37]. Therefore, we conducted comparative analyses between the ACS2N group and the other two groups in this study.

Our research demonstrates that 5-hmC signals are enriched in promoter, exon, UTR, and TTS regions. Additionally, through comparative analysis between the ACS2N group and the other two groups, we screened a total of 399 differential gene loci and conducted GO and KEGG enrichment analyses on them. GO analysis revealed significant enrichment of differentially expressed genes in multiple biological processes and molecular functional areas closely related to neutrophil functions. Specifically, these genes involve the regulation of viral life cycle, cellular response to cAMP, cAMP binding, and B cell differentiation, among other key biological processes. KEGG pathway enrichment analysis also highlighted that these differential genes mainly enriched in inflammation, immunity, and neutrophil-related signaling pathways. Numerous studies have reported that neutrophil-to-lymphocyte ratio (NLR) is typically elevated in peripheral blood of COVID-19 patients, especially in severe and critical cases [1, 2]. This increased NLR is closely associated with disease severity and prognosis, suggesting a significant role of neutrophils in the inflammatory response to COVID-19. Furthermore, neutrophils are closely related to the occurrence and development of ACS, and leukocytosis is widely suspected to play a causal role in ACS. Among leukocytes, neutrophil count is considered an independent prognostic factor for ACS patients, directly correlated with infarct size and left ventricular ejection fraction decrease after percutaneous coronary intervention or coronary artery bypass surgery[38]. Cytokine storm severity is a major factor involved in extrapulmonary organ damage in COVID-19[39]. Activation of the complement pathway and excessive release of cytokines such as interleukin-6 promote microthrombosis, leading to myocardial damage. In experimental models, complement pathway inhibition appears to have therapeutic effects against SARS-CoV and SARS-CoV-2 infections[40, 41].

Given the critical roles of inflammation and immunity in COVID-19 and ACS, we attempted to explore the relationship between differentially expressed genes and neutrophils in the three groups of ACS patients. The results showed a significantly higher proportion of neutrophils in ACS2N patients compared to the other two groups, consistent with our previous findings. It is inferred that these differentially expressed genes may play a crucial role in the occurrence and development of ACS by enhancing the activity and function of neutrophils.

Furthermore, we performed protein-protein interaction (PPI) network analysis on genes related to neutrophil signaling pathways. Through this strategy, we identified the PDE4D and PDE4B genes within the PDE family. Our research data shows that these two genes are upregulated in ACS patients and positively correlated with neutrophil activity. To further validate these findings, we analyzed external single-cell RNA sequencing datasets, confirming the expression patterns of PDE4D and PDE4B in neutrophils.

Phosphodiesterases (PDEs) influence intracellular signal cascades by mediating the hydrolysis of the second messengers cAMP and cyclic guanosine monophosphate (cGMP), thereby regulating various pathological and physiological processes[42]. PDE4 is the largest subclass of the PDE family, and PDE4 inhibitors increase cAMP levels by blocking PDE4 enzyme activity, thereby reducing the production of pro-inflammatory cytokines and inhibiting inflammatory responses. Therefore, PDE4 inhibitors have been widely developed for the treatment of various inflammatory diseases such as asthma, chronic obstructive pulmonary disease, and plaque psoriasis[42]. The team led by Wang Miao[43] first reported that phosphodiesterase 4B (PDE4B) plays a critical role in myocardial ischemia-reperfusion (MI/R) injury by mediating neutrophil inflammatory responses and microcirculatory disorders, which not only reduces myocardial cell death but also improves coronary microvascular obstruction, providing dual cardiac protective effects.

There have been no literature reports on PDE4D expression in ACS patients. Research by the team led by Zhong Nanshan[44] demonstrated that the E protein of SARS-CoV-2 upregulates PDE4D expression in airway epithelial cells, activating Toll-like receptors (TLR) 2/4 and downstream c-Jun N-terminal kinase (JNK) signaling, leading to increased intracellular chloride ion concentration ([Cl−]i). This elevation of [Cl−]i exacerbates airway inflammation by promoting serum/glucocorticoid regulated kinase 1 (SGK1) phosphorylation. Additionally, blocking SGK1 or PDE4 can alleviate E protein-induced intense inflammatory responses.

Previous studies have shown that increased PDE4D expression promotes vascular smooth muscle proliferation, migration, and exacerbates local inflammatory responses at vascular damage sites, thereby promoting the formation of atherosclerosis and increasing the instability of vascular plaques[45]. We hypothesize that high expression of PDE4D in neutrophils may promote inflammatory reactions by activating cAMP related signaling pathways, leading to poor prognosis in ACS. Therefore, we believe that COVID-19 induces neutrophil aggregation, upregulates PDE4D expression, activates cAMP-related signaling pathways, and promotes inflammatory processes, thereby affecting the occurrence and development of ACS. This provides a new research direction for the impact of COVID-19 on ACS.

However, our study has some limitations. Firstly, due to the limited sample size, even though certain trends were observed, the statistical significance and generalizability of the study results may be compromised. Secondly, the absence of basic experimental validation introduces uncertainty in the biological mechanistic explanations of our findings. Therefore, despite of the preliminary evidence regarding treatment effects, we emphasize the need for further validation and in-depth investigation for interpreting the results. Future research should focus on increasing sample sizes and conducting basic experimental validations to confirm and enhance the reliability and applicability of our findings.

## Conclusions

This study is the first to utilize 5hmC sequencing technology to reveal significant differences in 5hmC levels between ACS2N patients and ACS patients, indicating that 5hmC may play a crucial role in disease progression. Meanwhile, no significant differences in 5hmC levels were observed between ACS2W patients and ACS patients, suggesting that the impact of COVID-19 infection on ACS patients two months later is relatively minor. Further analysis also revealed that the proportion of neutrophils was significantly higher in ACS2N patients compared to the other two groups. This finding suggests that COVID-19 infection may exacerbate the onset and progression of ACS by enhancing neutrophil activity. Additionally, a detailed analysis of differentially expressed genes, with a focus on PDE4D, found that PDE4D expression is positively correlated with neutrophil activity, and PDE4D expression was significantly elevated in ACS2N patients. Based on these findings, we hypothesize that COVID-19 infection may trigger upregulation of PDE4D gene expression, which in turn promotes neutrophil activation, potentially contributing to poor prognosis in ACS following COVID-19 infection.

## Data Availability

All data included in this study are available upon request by contact with the corresponding author.

## Abbreviations

COVID19: Corona Virus Disease 2019
ACS: Acute Coronary Syndrome
ACS2N: ACS within 2 months after COVID-19 infection group
ACS2W: ACS more than 2 months after COVID-19 infection group
cfDNA: Cell-free DNA
5hmC: 5-Hydroxymethylcytosine
5mC: 5-Methylcytosine
PDE4D: phosphodiesterase 4D
RAAS: renin-angiotensin-aldosterone system
STEMI: ST segment Elevation Myocardial Infarction
MACE: major adverse cardiovascular events
HR: Hazard Ratio
BMI: body mass index
hsCRP: hypersensitive c-reactive protein
Cr: creatinine
AST: aspartate aminotransferase
ALT: alanine aminotransferase
TG: triglyceride
CK-MB: creatine kinase-MB
LVEF: left ventricular ejection fractions
TnT: troponin T

## Data availability statement

The datasets supporting the conclusions of this article are included within the article and its additional files. All other datasets used and analyzed during the current study are available from the corresponding author on reasonable request.

## Ethics statement

The study was conducted according to the guidelines of the Helsinki Declaration and was approved by the Ethics Committee of Peking University Third Hospital. Written informed consent was obtained from all participants.

## Consent for publication

Not applicable.

## Author contributions

Long Chen and Lijie Sun conceived the research and designed the experiments. Zhongheng Li, Ruining Li, Dan Li and Yishuo Liu recruited patients, collected blood samples, and registered clinical information. Hangyu Chen and Lei Zhang participated in library construction and 5hmC sequencing. Maimaitiyasen Duolikun conceptualized and oversaw the study, analyzed data. Zhongheng Li and Maimaitiyasen Duolikun wrote the manuscript. Lijie Sun, Hangyu Chen and Long Chen revised the manuscript. All authors were responsible for conceptual design, data analysis, manuscript writing, and submission.

## Acknowledgments

We would like to acknowledge the essential contributions of all staffs and students who participated in this work.

